# Indian Rare Disease Stakeholder Mapping

**DOI:** 10.1101/2024.07.04.24309947

**Authors:** Mohua Chakraborty Choudhury, Jerry Philip George, Prashanth N Srinivas

**Affiliations:** DST Center for Policy Research Indian Institute of Science, Bengaluru, India; Institute of Public Health, Bengaluru, India; Sree Chitra Institute for Medical Sciences and Technology, Thiruvananthapuram, India

**Keywords:** RD policy, stakeholder analysis, health policy, public health, Stakeholder, stakeholder mapping

## Abstract

Rare diseases (RD) aren’t rare collectively, affecting around 300 million people globally and 96 million in India. In low- and middle-income countries like India, policies addressing these diseases have only recently been enacted. In 2021, India launched its first functional RD policy. This study comprehensively maps all stakeholders in the RD ecosystem in India to understand their power positions, influence, and needs, thereby enabling better implementation strategies for the RD policy. We conducted in-depth interviews with various stakeholders to understand their perspectives and supplemented the study with media analysis to reach those who did not respond to interview invitations. Our findings suggest a lack of awareness and knowledge about RDs among healthcare professionals who do not specialize in RDs. Encouraging and formalizing the involvement of RD patient organizations in policy-making is crucial due to their high knowledge, interest, and constructive critical capabilities despite their low power. Another important stakeholder group, local companies, can drive innovation and make treatments accessible for RDs but have much lower power than multinational companies, potentially leading to policies that do not favor local needs.

## 1. Introduction

Severe debilitating medical conditions that affect only a small portion of the population are classified as rare diseases (RD). Despite the low prevalence rate, RDs collectively affect a substantial population of around 300 million individuals worldwide at any time (1) and pose a significant challenge to the healthcare system of any country. They can also have a severe impact on the lives of patients and their caregivers. In India about 96 million patients are affected by RDs (2) (3) In the last decade there was very little support from the government for these patients, and RDs did not feature in the country’s health policy agenda. However, driven by patient advocacy movements in 2017 Ministry of Health and Family Welfare government of India launched the country’s first National Policy for RDs which was put in abeyance in 2018, and a revised version was adopted in 2021 (4). The vision of the National Policy for RDs in India is to ensure early diagnosis, affordable treatment, comprehensive support systems, and collaborative research to improve the lives of RD individuals’ policy (5). The policy faced mixed reviews from different stakeholders. However, most appreciated it as a move in the right direction (6).

The biggest task that lies ahead is the implementation of the vision laid out in the policy. In general, stakeholders play a crucial role in the RD ecosystem due to the unique challenges posed by these conditions with the varying definitions and limited prevalence rates, it becomes imperative for stakeholders, including patients, caregivers, healthcare providers, researchers, advocacy groups, and policymakers, to collaborate and work together to address the complexities of RDs. By pooling their knowledge and resources, stakeholders can raise awareness about RDs, facilitate early diagnosis, and ensure access to appropriate healthcare services (7).

Thus it is crucial to identify all the stakeholders in the RD ecosystem, and understand their position, power, role, and influence (8). So that a collaborative approach can be adopted to bring all the stakeholders on the same platform and work towards implementation of the policy. In this study, we attempt to identify all major stakeholder groups in India and understand their knowledge, power, position, and interest in the RD ecosystem and NPRD. This is the first study to map RD stakeholders in a low- and middle-income country (LMIC), making it crucial for similar efforts in other LMICs.

## 2. Methodology

We used qualitative research methodology that involved conducting in-depth interviews and media analysis. In our exploratory mapping study, we aimed to identify major groups of stakeholders and contacted as many individuals as possible within each group to achieve content saturation. However, it was not practically feasible to identify all possible stakeholders in each group. Despite efforts, some groups had insufficient response, requiring analysis with limited data. To supplement this, additional media analysis was conducted for groups with fewer respondents. The different steps involved in the study is laid out in Fig 1 and each step is described below:

**Figure 1.**
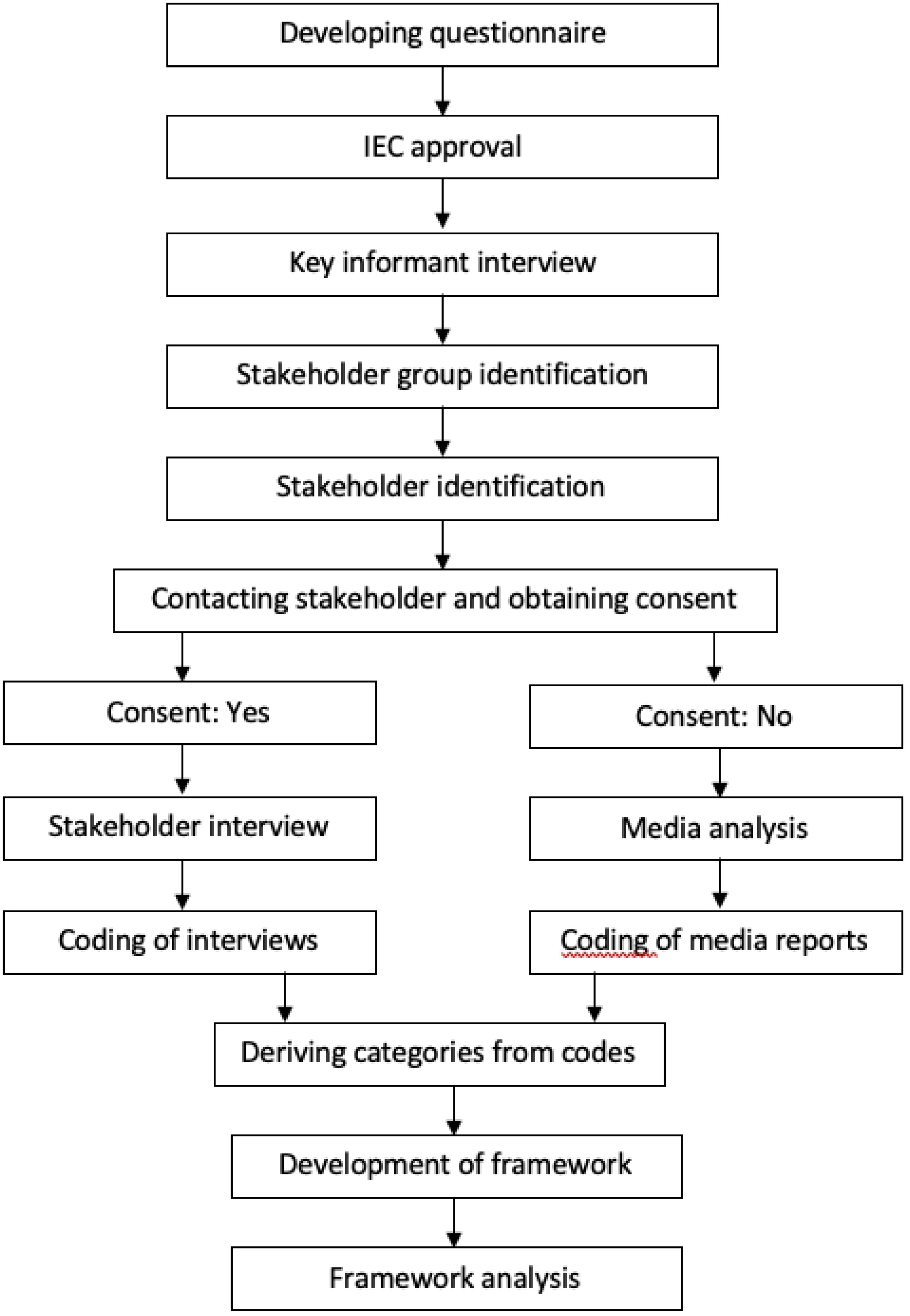
A flowchart showing the key steps of methodology.

### 2.1 Developing questionnaire

The questionnaire for this study was collaboratively developed by the corresponding author, MCC and PNS, a co-author and mentor in this study. Drawing upon MCC’s knowledge and research experience of the RD ecosystem over the past six years, as well as PNS’s extensive expertise in the field of public health, we designed the questionnaire to capture comprehensive insights from the stakeholders for the RD ecosystem. This enabled the development of a semi-structured questionnaire as it would help to provide a balance between flexibility and structure, allowing for in-depth exploration of research topics while ensuring consistency across participants.

### 2.2 Ethics, Consent, and Permission

To ensure ethical conduct, the study received approval from the Institutional Ethics Committee of the Institute of Public Health for study number IEC-FR/01/2021 on 04/02/2021. The potential interviewees were provided with an information sheet containing information about the researchers’ affiliation, the source of funding, the study objectives, and the investigators’ plan to publish the findings in an academic journal without seeking their approval. Participants were required to respond positively to an interview invitation sent by email and sign an informed consent form to indicate their willingness to participate in the interviews. While confidentiality was assured, we sought permission to use specific information from the interviews in the intended publication. Lastly, participants were informed that they would not receive any compensation for participating in the study.

### 2.3 Key informant interviews

A series of four key informant interviews were conducted in order to facilitate contact with various stakeholders within the RD ecosystem. A key informant was identified as a person who is knowledgeable about the RD ecosystem and has ideas about different stakeholder groups but is not inherently a stakeholder. These interviews served as a valuable means of identifying and categorizing stakeholders into relevant groups.

### 2.4 Stakeholder identification and categorization

Through the initial key informant interviews, background literature survey and authors previous experiences of Indian RD ecosystem research, the investigator was able to identify a total of 10 distinct stakeholder groups within the RD ecosystem. These groups included doctors with specialization in RDs, general physicians, Indian and international organizations focused on RDs, patient organizations, think tanks, the research community, multinational corporations, policymakers and government, and allied health professionals. This comprehensive categorization process allowed for a more systematic analysis of the different types of stakeholders involved in RDs. We contacted several stakeholders across these groups but were finally able to get interviews from a total of 33 stakeholders. After receiving consent from eligible participants, in-depth interviews were carried out to further explore the perspectives and experiences of stakeholders within the RD ecosystem.

### 2.5 Stakeholder interviews

Eligible stakeholders who expressed their consent to participate were subsequently invited to participate in an in-depth semi-structured interview via either online or offline modes, based on their preference and convenience. The interviews were conducted between May 2021-July2022. Interviews were conducted via video conference and recorded with participants’ prior consent. Written and oral consent were obtained after detailed explanation of the informed consent form (ICF). Participants were given sufficient time to review the ICF and ask questions. Oral consent was reconfirmed at the start of each interview. However, it is worth noting that despite efforts to engage all identified stakeholders, a few were unable to participate due to no response to the invitation emails or scheduling conflicts. Nonetheless, the investigator was able to conduct interviews with a substantial number of stakeholders in different categories to get an understanding of the stakeholder perspective in each category. This is an exploratory study and it gives a reflection of perspectives of different stakeholder categories and does not intend to provide a conclusive inference of perspectives of any group.

### 2.6 Media analysis

Despite the efforts to engage with all categories of stakeholders, some groups such as MNCs, policy makers, government officials, and the judiciary had limited participation in in-depth interviews. As a result, an alternative data collection method, media analysis, was implemented. To conduct this analysis, relevant news articles related to RDs and the respective stakeholders were searched using specific keywords on Google, covering the period from March 2017 to September 2022. Results from the first five pages of the search were considered, and the articles were then subjected to qualitative analysis as a substitute for in-depth interviews (Table 4).

### 2.7 Qualitative analysis of stakeholder interviews

The transcriptions of the in-depth interviews were generated using Otter AI, and the qualitative analysis was performed using QDA Miner 6 for Windows. Initially, an inductive coding approach was utilized to code at least one interview from each stakeholder group. This coding enabled the derivation of codes and categories that were subsequently used for the deductive coding of the remaining interviews. By incorporating both inductive and deductive coding methods, a hybrid approach was utilized for the qualitative analysis. However, in some cases, new codes were added while analyzing these sets of interviews aswell.

### 2.7 Qualitative analysis of media articles

The media analysis also involved a qualitative analysis of the relevant news articles. In this approach, the entire articles were used as a substitute for in-depth interview transcripts. Any quotes or reference to a stakeholder was extracted from their respective perspective. Deductive coding was then applied to these articles, based on the codes and categories that were formed from the in-depth interview analysis. By utilizing this approach, it was possible to further explore the themes and insights related to the stakeholder groups that had limited participation in the in-depth interviews. The findings from the media analysis were also integrated with those from the in-depth interviews to provide a more comprehensive understanding of the RD ecosystem and the stakeholders involved.

### 2.8 Development of framework

In order to conduct further analysis of stakeholders in the context of the study, the stakeholder analysis framework originally developed by Balane MA et al. (9) was adapted and customized as necessary. After thorough consideration, it was determined that this framework was indeed appropriate for the study, with only minor changes required to fully align it with the specific context of the RD ecosystem being analyzed. Within this adapted framework, stakeholders were assessed and evaluated based on four key parameters: knowledge, interest, power, and position. This approach provided a comprehensive and structured means of analyzing and categorizing stakeholders, allowing for a more systematic understanding of their roles, motivations, and potential impact within the ecosystem.

### 2.9 Framework analysis

The stakeholder framework by Balane MA et al was adopted and modified to suit the present study (Balane et al., 2020). The stakeholder analysis framework utilized a four-point scoring system for evaluating each of the four stakeholders’ characteristics: knowledge, interest, power, and position. The scores obtained were used to assess these characteristics in each stakeholder group, enabling a comprehensive understanding of their roles and perspectives. The scoring process was based on the codes and categories derived from the qualitative interviews, allowing for a more nuanced analysis of the stakeholders. The qualitative analysis of the study was used to score stakeholders based on knowledge, interest, power, and position on a four-point scale ranging from 0-3. To fit the context of the study, the definition and interpretation of the value scale were also modified from the Balane et al. framework. The ‘knowledge’ and ‘interest’ components in the modified framework are fairly similar to the components in the original framework. In power, an additional entity of ‘no power’ in the value scale interpretation was added as a few stakeholders in the study had no influence over policymaking. Similarly, in position, value scale definition and interpretation were modified so as to ascertain the position that each of the stakeholders took in the study. Table 1 gives a detailed description of the framework and lists the domains used in each characteristic.

**Table 1.** Modified stakeholder analysis framework The original framework adopted from Bayne et. al. was modified to suit the RD ecosystem in India.

| Modified SH Analysis Framework Adopted from Balane MA, Palafox B, Palileo-Villanueva LM, et al |  |  |  |  |
| --- | --- | --- | --- | --- |
| SH Characteristics | Characteristic definition | Domain | Value scale definition | Value scale interpretation |
| Knowledge | SH's knowledge and understanding of the RDs and RD policy | •Knowledge of RDs<br>•Knowledge of RD policies<br>•Knowledge of RD management<br>•Understanding of Indian RD ecosystem | 0-No knowledge in the listed domains | 0—No knowledge |
|  |  |  | 1-Knowledge in 1 listed domains | 1—Limited knowledge |
|  |  |  | 2-Knowledge in 2 listed domains | 2—General Knowledge |
|  |  |  | 3-Knowledge in 3 or more listed domains | 3—Extensive knowledge |
| Interest | SH's interest in RD policy development, implementation and contribution to RD ecosystem | •Interest in RD policy development<br>•Interest in RD policy implementation<br>•Contribution to RD ecosystem | 0-No interest in the listed domains | 0—No Interest |
|  |  |  | 1-Interest in atleast 1 domain | 1—Limited Interest |
|  |  |  | 2-Interest in 2 listed domains | 2—General Interest |
|  |  |  | 3-Interest in 3 listed domains | 3—High Interest |
| Power | The ability of the SH to influence RD policy development and implementation | •Political authority<br>•Financial authority<br>•Technical (Domain) expertise<br>•Leadership | 0-Stakeholder possesses and has no control over any of the listed domains | 0—No power |
|  |  |  | 1-Stakeholder possesses and has control over use of one to two domains of power, low potential to affect policy implementation | 1---Low power |
|  |  |  | 2-Stakeholder possesses and has control over use of two to three domains of power, has moderate potential to affect policy implementation | 2—Medium power |
|  |  |  | 3-Stakeholder possesses and has control over use of three to four domains of power, has high potential to affect policy implementation | 3—High power |
| Position | Whether the SH supports, opposes or is neutral about policy implementation | A.degree of support or opposition to policy expressed through use of potential power (sources of power)<br>1. Neutral | 0- Opponent: A4 and B1 or B2 | 0---Opponent: Stakeholder uses potential power to moderately acting against policy implementation |
|  | on | 2. Supporter<br>3. Moderate Supporter<br>4. Opponent<br>B.Actions taken to demonstrate support or opposition to policy<br>1. No actions taken<br>2. Actions taken | 1- Neutral: A1 and B1 | 1—Neutral: Stakeholder does not use potential power and does not act for or against policy implementation |
|  |  |  | 2- Moderate Supporter A3 and B1 or B2 | 2—Moderate Supporter: Stakeholder uses potential power to moderately act in support of policy implementation |
|  |  |  | 3- Supporter: A2 and B1 or B2 | 3—Supporter:Stakeholder uses potential power to strongly act in support of policy implementation |

Through these domains, we intended to capture the breadth of the characteristics of each stakeholder across all the listed domains. A stakeholder having characteristics in more than one domain is rated higher on the value scale. If a SH’s characteristics lie only in one domain it will be rated lower even if he has deep expertise in that domain. Thus the matrix does not aim to measure the depth of SHs in each domain because the focus of the matrix is to elucidate the overall understanding of each stakeholders about the RD ecosystem and not depth of knowledge in their respective domain. Also, the authors do not have expertise to assess depth of knowledge in each domain and the questionnaire was designed accordingly.

## Results

In-depth interviews served as the primary method for data extraction. The key informants were interviewed first, followed by the stakeholders.

### 3.1 Key informant interviews

Key informant interviews played a crucial role in identifying and defining the stakeholder categories by highlighting potential stakeholders within the RD (RD) ecosystem. The insights provided by the key informants proved invaluable in understanding the primary stakeholders and their specific roles and functions. Through their discernment, various stakeholders were identified, contributing to a comprehensive understanding of the RD ecosystem and laying the foundation for subsequent analysis and investigation.

### 3.2 Profile of stakeholders

A list of all potential stakeholders was created with information about their affiliations, and each stakeholder was contacted. Many participants spoke from personal experience rather than on behalf of their organizations. However, their viewpoints might be seen as a reflection of their experience and engagement with the respective organizations. A total of 33 in-depth interviews (IDI) were taken including participants from different categories. Stakeholders who did not respond to our invitation to participate in the interview even after several attempts till November 2022 were selected for media analysis. A summary of the profile of the participants is given in table 3.1.

### 3.3 Codes and Categories

The deduced codes were grouped into seven major categories described here.

(i) **Experience**: This category was formed out of codes that encompassed the personal and professional experience the stakeholders had in the RD ecosystem. It also included codes on contributions made towards advocacy, research, and activities related to RD.
(ii) **Perspectives /Knowledge:** This includes codes on knowledge of RDs, RD policies, diagnosis, treatment, clinical trials, the registry, and orphan drugs. It also included codes for understanding the Indian RD ecosystem and the drivers of change.
(iii) **Challenges**: The RD stakeholders faced challenges at multiple levels, including sociocultural, policy-level, research, infrastructure, healthcare access, treatment cost, inclusivity in policymaking, insurance, and healthcare worker competency. These were essentially captured into codes and helped in forming the category of challenges in the RD ecosystem in India.
(iv) **Recommendations:** Similarly, recommendations included codes on policy refining, social and emotional support, awareness improvement, funding, screening, and improving access to diagnosis and treatment.
(v) **Influence:** Influence on RD-related policies had codes classified into two categories: direct influence and indirect influence. Direct influence occurs when a stakeholder has a direct role in policymaking, whereas indirect influence occurs when a stakeholder exerts influence through other activities such as empowering PAGs or holding meetings, discussions, or conferences to foster a favorable policy environment.
(vi) **Alliance:** Another category that aided in identifying the networks that existed between various stakeholders was alliance. Codes in this category included knowledge about policy alliances and organizational alliances, such as those between industry, academia, the PAG, the government, and international organizations.
(vii) **Implementation:** In the implementation category interest, attitudes, challenges, and recommendations pertaining to the NPRD were captured.

The summary of categories and codes is given in Table 3.2

### 3.4 Media Analysis

Media analysis helped to compensate for underrepresentation in some stakeholder groups mainly in policymakers and government, judiciary and multinational companies. A total of 127 articles were extracted and reviewed from the timeline of 2017 to 2020. This presented an interesting view of a wide distribution of news articles containing RD coverage in India. Yearly distribution of newspaper reports covering news and stories related to RDs was low which showed the lack of media interest in this field. However, the coverage has shown some increase over time with the release of the first National RD Policy in 2017. Relevant quotes and information related to each of the above-mentioned characteristics for stakeholders in this group were captured and analyzed along with interview-data.

### 3.5 Stakeholder analysis and interpretation

Stakeholder analysis based on the Blane MA et al framework has 4 key stakeholder characteristics mainly knowledge, interest, power, and position. These characteristics were adopted and defined based on the context of the RD ecosystem in India. Knowledge refers to the stakeholder’s knowledge and understanding regarding RDs and their ecosystem in India. Interest refers to stakeholders’ interest in RD policy development, implementation, and contribution to the RD ecosystem. Power refers to the ability of the stakeholder to influence RD policy development and implementation. Position refers to the alignment of the stakeholders whether they support, oppose, or are neutral about policy implementation. The domains of each of the 4 characteristics are enlisted in Table 3

**Table 2:** Categories of Stakeholders, Abbreviated Codes, and Data Collection Methods. This table provides an overview of the stakeholder categories included in the study, along with their corresponding abbreviated codes and methods of data collection.

| Sl. No. | Stakeholder group | Code | Method of data collection |
| --- | --- | --- | --- |
| 1 | Doctors (non-RD specialists) | NRDS | In-depth interviews |
| 2 | Doctors (RD specialists) | RDS | In-depth interviews |
| 3 | International patient organizations | INTO | In-depth interviews |
| 4 | Indian Orphan Medicinal Product Organization | LOCO | In-depth interviews |
| 5 | Indian patient organizations | PATO | In-depth interviews |
| 6 | Think tanks | TTNK | In-depth interviews |
| 7 | Research community | RCOM | In-depth interviews |
| 8 | Multinational companies | MNCO | In-depth interviews & media analysis |
| 9 | Policymakers and government | PMKR | In-depth interviews & media analysis |
| 10 | Allied health professionals | AHPR | In-depth interviews |

**Table 3:** Categories of Classified Interview Codes This table presents an overview of the categories and codes derived from the qualitative analysis of in-depth interviews and media analysis. The framework component contributed by each stakeholder category is displayed in the right column.

| Sl. No. | Category | Description of the category | Framework component |
| --- | --- | --- | --- |
| 1 | Experience with RD | Included codes on personal and professional experience with RD and contributions made in advocacy, research and activities related to RD | Interest |
| 2 | Knowledge on RDs and related policies | Included codes on knowledge of RDs, RD policies, diagnosis, treatment, clinical trials, registry and orphan drugs. It also includes codes on understanding of the Indian RD ecosystem and the drivers of change. | Knowledge |
| 3 | Challenges in RD ecosystem in India | Includes codes on challenges at various levels – sociocultural, policies, research, infrastructure, healthcare access, cost of treatment, inclusivity in policymaking, insurance and competency of health care workers | Knowledge |
| 4 | Recommendations | Included codes on policy refining, social and emotional support, awareness improvement, funding, screening, improving access to diagnosis and treatment | Knowledge, Interest |
| 5 | Influence on RD related policies | Codes are divided into 2; direct and indirect influence. Direct influence is when a stakeholder has direct role in policymaking and indirect influence represents influence via other activities such as empowering PAGs, meetings/discussions/ conferences held to make favourable environment for policy. How they | Power |
| 6 | Alliance | Includes codes about knowledge on alliance between policies and on alliance between organizations such as alliance between industry, academia, PAG, govt and international organizations | Power, Knowledge |
| 7 | Implementation | Includes codes on role in implementation, interest, attitude, challenges and recommendations pertaining to NPRD | Interest , Power , Position |

**Table 4.** Keywords and the number of articles reviewed in media analysis. This table presents a list of keywords used for media analysis along with the corresponding number of articles reviewed for each keyword by year. The number of articles reviewed may vary depending on the availability and relevance of articles related to each keyword.

| Sl.No. | Key word | Number of relevant articles |  |  |  |  |  |
| --- | --- | --- | --- | --- | --- | --- | --- |
|  |  | 2017 | 2018 | 2019 | 2020 | 2021 | 2022 |
| 1 | MOHFW RD policy India | 3 | 1 | 2 | 6 | 4 | 5 |
| 2 | Niti Aayog RD policy India | 2 | 0 | 2 | 5 | 4 | 5 |
| 3 | Ministry of finance RD policy India | 6 | 3 | 2 | 1 | 3 | 4 |
| 4 | CDSCO RD policy India | 1 | 1 | 2 | 1 | 3 | 4 |
| 5 | DCGI RD policy India | 0 | 2 | 0 | 3 | 3 | 3 |
| 6 | Court RD policy India | 2 | 4 | 2 | 4 | 3 | 4 |
| 7 | Karnataka RD policy India | 2 | 1 | 0 | 4 | 3 | 4 |
| 8 | Kerala RD policy India | 3 | 1 | 0 | 4 | 3 | 3 |

#### 3.5.1 Distribution of characteristics: knowledge, interest, position, and power within each stakeholder category

##### Knowledge

Knowledge was assessed from 4 key domains namely knowledge of RDs, knowledge of RD policies, knowledge of RD management, and understanding of the Indian RD ecosystem which were derived from the codes of interviews.

Figure 2, which depicts a Box and Whisker plot, reveals notable variations in knowledge levels among different stakeholders in the study. AHPR (2.4), NRDS (1.75), and RCOM (2.6) exhibit significant intra group variation, indicating a wide range of knowledge scores within these groups. Conversely, the remaining groups predominantly display a similar high score in knowledge, suggesting a more homogeneous distribution within these groups.

**Figure 2.**
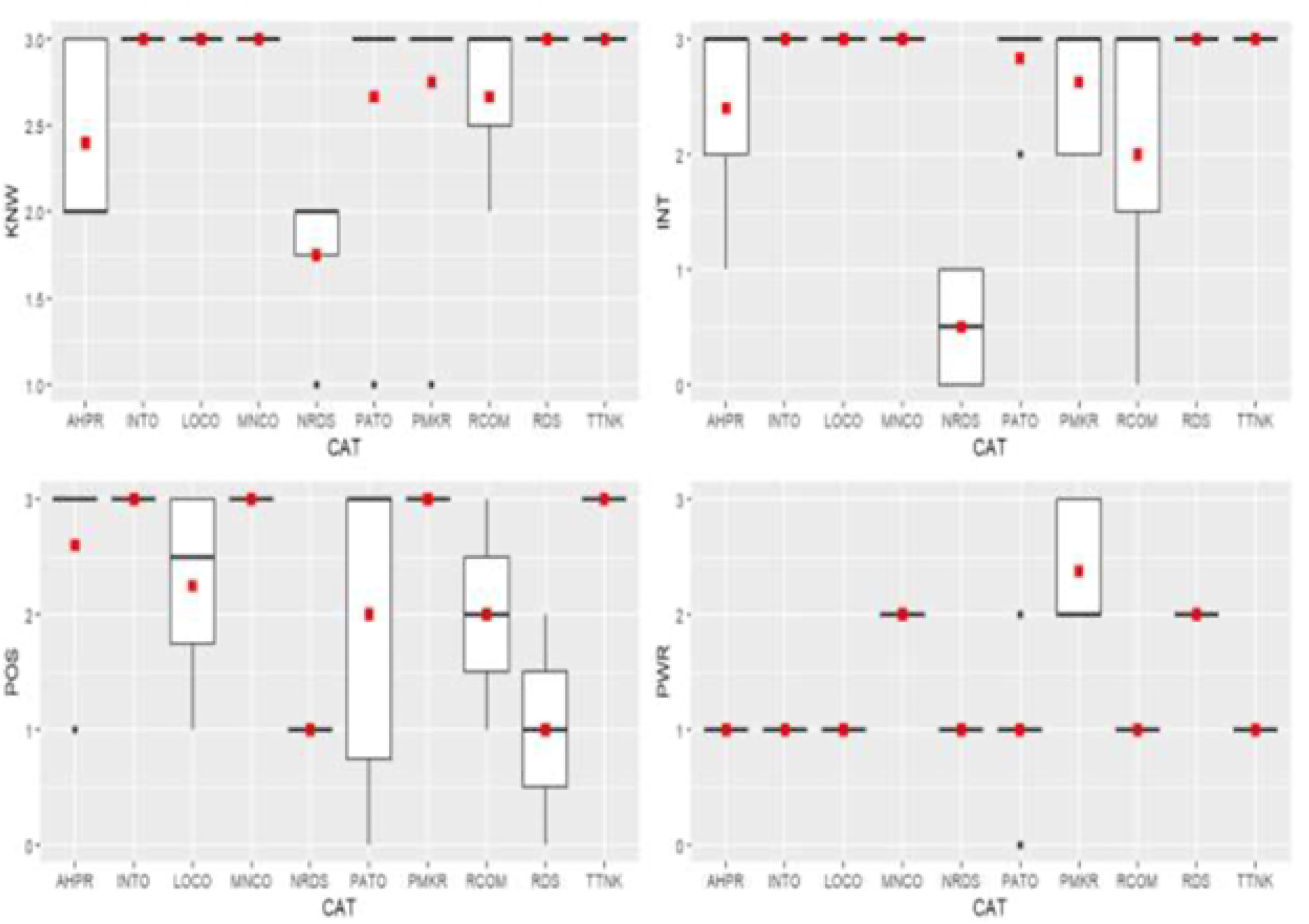
Box and Whisker Plots illustrating the distribution of knowledge (KNW), interest (INT), position (POS), and power (PWR) among each stakeholder category (CAT). The distribution of knowledge, interest, position, and power scores among the stakeholders included in the study. The whiskers extend to the minimum and maximum values, excluding outliers represented as individual data points beyond the whiskers. The red dots indicate mean scores obtained by each stakeholder category.

**Figure 3.**
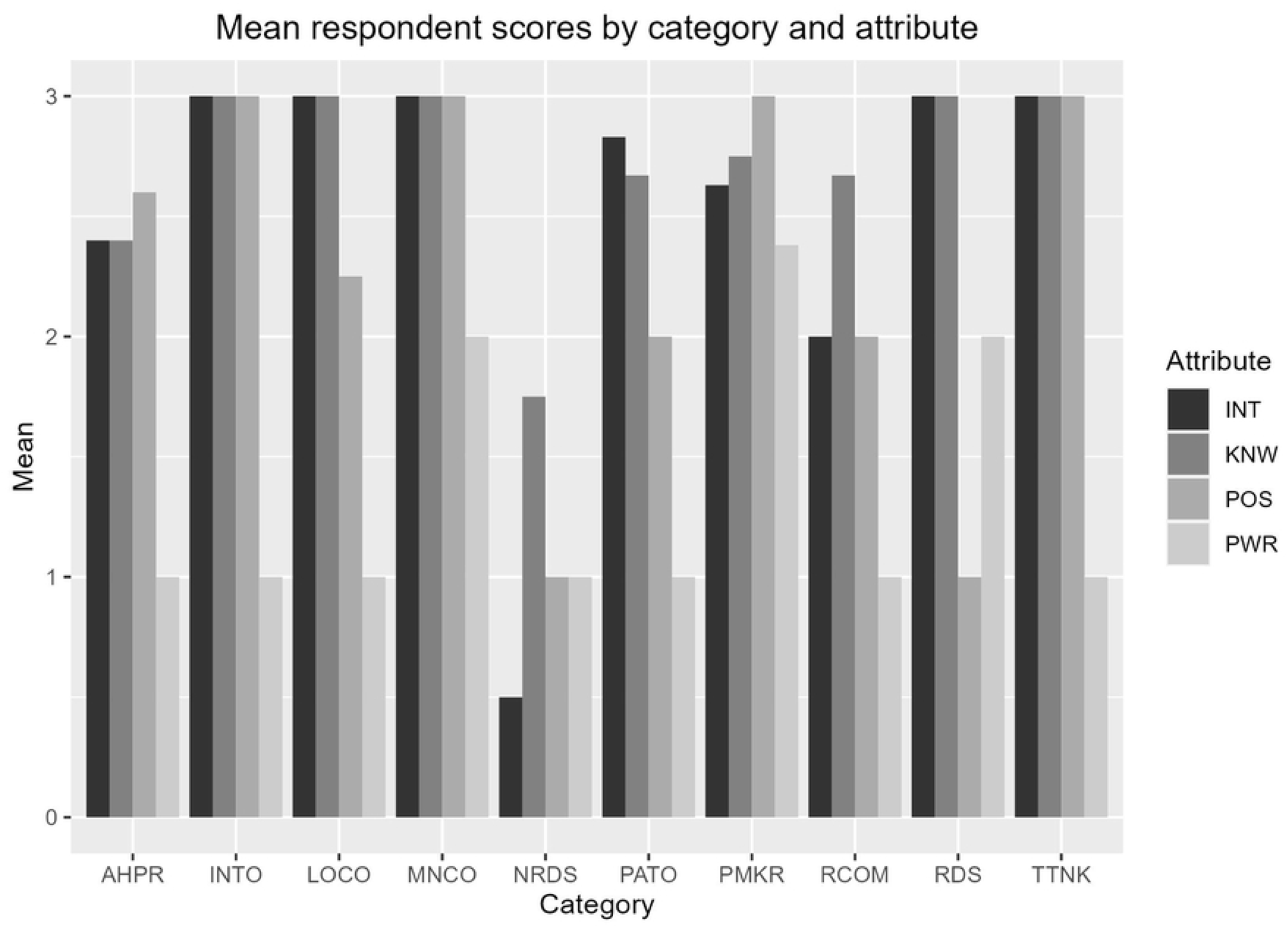
Grouped column chart of mean respondent scores by stakeholder category and characteristics (Knowledge, Interest, Power, and Position).

Extensive knowledge was observed in RDS (2.7), INTO (3.0), LOCO (3.0), PATO (2.6), TTNK (3), RCOM (2.6), MNCO (3), and PMKR (2.7). A limited /general knowledge was seen in NRDS (1.8) who are not specialized in RDs as most of them have not encountered an RD patient and are not aware of various diagnostic possibilities as well.

Stakeholders who possess limited or general knowledge in one domain may have extensive knowledge in other domains. RDS has extensive knowledge of disease, diagnosis, treatment, and care. However, all of them might not be well acquainted with RD policies. The extensive knowledge possessed by RDS arises from their experience as treating physicians. For example, a specialist we interviewed who specializes in rare skin diseases revealed that he is one of the very few clinicians in India treating the RD Epidermolysis bullosa. Patients from many states visit this SH. Similar to this, another expert who not only practices medicine but also serves in a variety of other roles, such as those related to policy making, has a thorough knowledge of the ecosystem in general as well as the clinical and academic aspects of RDs. PATOs actively engage in various activities such as campaigning, fund generation, and mobilizing various actors under the same umbrella to gain a collective benefit or be a strong voice to speak for their needs. Many of the actors in these organizations are patients or their caretakers, who have first-hand knowledge of the disease and its sufferings. Their voice is the most dominant among all the other actors, and therefore they are well networked with multiple stakeholders. For instance, it was only after multiple lawsuits from some of the PATOs that the judiciary had to intervene to order the government to make a new policy, which resulted in the National RD Policy of 2017. INTOs offer support in many roles, such as providing technical expertise, exposing the Indian counterparts to the policies of other countries, mobilizing funds, and helping to represent India on various international platforms pertaining to RDs.

LOCO, RCOM, MNCO, and TTNK possess extensive technical knowledge in their own respective areas of interest. RCOM contributes to evidence generation. TTNK helps in organizing existing evidence to support policy making. MNC and LOCO are commercial players who have their own research wings that aid in creating new products that are affordable to Indian patients. For instance, we interviewed a company that specializes in nutritive products specifically designed for RD patients. They have devoted years to research for developing affordable products specific to particular RDs in India which were previously imported and were exorbitantly priced.

NRDS and APHR have limited knowledge about the clinical aspects of RDs including screening and diagnosis. Their experience with RDs is low both in terms of academic exposure to RDs and seeing RD patients. A few NRDS at primary health centers revealed that they did not come across any patients during their practice.

##### Interest

Interest was assessed based on 3 domains, namely, interest in RD policy development, interest in RD policy implementation, and contribution to the RD ecosystem. As seen in Fig 2 High interest was observed among RDS (3.0), INTO (3.0), LOCO (3.0), PATO (2.9), TTNK (3.0), and MNCO (3.0). A general/limited interest was observed in NRDS (0.5) and AHPR (2.4)

Figure 2b, further illustrates the variations in interest levels among different stakeholders in the study. It is observed that RCOM exhibits significant intra group variation, followed by AHPR, NRDS, and PMKR, indicating a wide range of interest scores within these groups. Conversely, the majority of stakeholders in the remaining groups demonstrate a similar high score in interest, suggesting a more consistent distribution within these groups.

TTNK and RCOM are more inclined towards the academic aspects and therefore have an interest in the policymaking process which might contribute to research activities in the ecosystem. On the other hand, MNCO’s interest in RD policymaking is driven by their own business interests. PMKRs showed a wide disparity in interest within the group in policy development and implementation. A limited interest was shown by a few SHs in this group as competing priorities in a resource-constrained setup limit their ability to address the pertinent issues related to RDs significantly.

##### Power

According to Figure 2, the power levels among different stakeholder groups in the study exhibit variations. Notably, the PMKR (2.4) group stands out with wide intragroup variation and high power scores (mean 2.4). Additionally, the MNCO (2.0) and RDS (2.0) groups demonstrate medium power levels. In contrast, the remaining stakeholders across different groups display low power scores (1.0).

Power was also assessed based on 4 domains, namely, political authority, financial authority, technical (domain) expertise, and leadership. Power was concentrated more in PMKR followed by MNCO and RDS. PMKRs had the upper hand in all 4 domains. MNCO possessed a medium power as they had the upper hand only in 2 domains - financial authority and technical domain expertise and can influence policymakers to a certain extent. On the other hand, RDS had a medium power to influence policy making and implementation as most of the specialists in the study are involved in the policymaking process through participation in various task forces. These stakeholders have a thorough understanding of the RD ecosystem and hence their contributions through various task forces can make meaningful changes. They have first-hand experience and a better understanding of the patient journey and hence can serve as a bridge between the patients and the policymakers. In this way, a balance is obtained in power dynamics where stakeholders of lower power can voice their recommendations through stakeholders of higher power. All other stakeholders namely NRDS, INTO, LOCO, PATO, TTNK and RCOM possessed low power. These stakeholders had an upper hand only in one of the four domains. They are well-wishers of the policy and are generally supporters of the existing state of affairs in the ecosystem. Most of these stakeholders have an upper hand in technical expertise in the respective areas they engage in.

##### Position

Figure 2c, depicted as a Box and Whisker plot, highlights the variations in positions among different stakeholders in the study. The plot reveals wide intragroup variation in the LOCO (1.0), PATO (2.0), RCOM (2.0), and RDS (2.3) groups, suggesting a diverse range of positions within these groups. On the other hand, the NRDS (1.0) group is characterized by a neutral position, indicating a lack of clear alignment with any specific stance. Within the PATO and RDS groups, a few stakeholders are shown to oppose the policy, indicating dissenting positions. In contrast, the remaining groups are predominantly identified as supporters of the policy.

Position was assessed based on 2 key domains: the degree of support or opposition to policy expressed through the use of potential power and actions taken to demonstrate support or opposition to the policy. Position refers to the alignment of the stakeholder whether they support, oppose, or are neutral about policy implementation. NRDS took a neutral position as they had a limited understanding of the RD ecosystem or the activities pertaining to the same. A few SHs in PATO and one member of RDS opposed the policy owing to a few limitations of the policy. These stakeholders have an extensive first-hand understanding of RDs and the policy and have been proactive in this space. They are aware of the limitations of the policy as well as have an understanding of how policies around the world pertaining to RDs are functioning. All other stakeholders namely NRDS, INTO, LOCO, TTNK, and RCOM were supporters of the policy. These stakeholders are in favor of the existing norms in the policy and are optimistic in general about the present state of affairs in the ecosystem.

#### 3.5.2. Comparative analysis of mean respondent scores across the stakeholder categories for each characteristic

The grouped column chart presents the scores of respondents within each stakeholder category for the four characteristics: knowledge, interest, power, and position. Each column represents the score of a specific stakeholder category for a particular characteristic. The x-axis represents the stakeholder categories, while the y-axis represents the scores. This presents a comparative analysis in each category between different stakeholder groups.

Knowledge and interest are high among most stakeholders. Different stakeholders take different positions. However, major variation is seen in Power between different stakeholders.

The provided scatter plot Figure 4 presents a comprehensive summary of the characteristics exhibited by the 10 stakeholders under investigation. NRDS stakeholders were found to possess limited knowledge, limited interest, low power, and maintained a neutral position. On the other hand, RDS stakeholders demonstrated high knowledge, high interest, medium power, and displayed varying positions. INTO stakeholders exhibited high knowledge, high interest, low power, and actively supported policy initiatives. Similarly, LOCO stakeholders showcased high knowledge, high interest, low power, and were supporters of policy. PATO stakeholders were characterized by high knowledge, high interest, low power, and displayed varying positions. TTNK stakeholders exhibited high knowledge, high interest, low power, and actively supported the policy. RCOM stakeholders possessed high knowledge, high interest, low power, and were moderate supporters. MNCO stakeholders displayed high knowledge, high interest, medium power, and were supporters. PMKR stakeholders demonstrated high knowledge, varying interests, high power, and actively supported the policy. Lastly, APHR stakeholders had varying levels of knowledge, varying interest, low power, and occupied varying positions.

**Figure 4.**
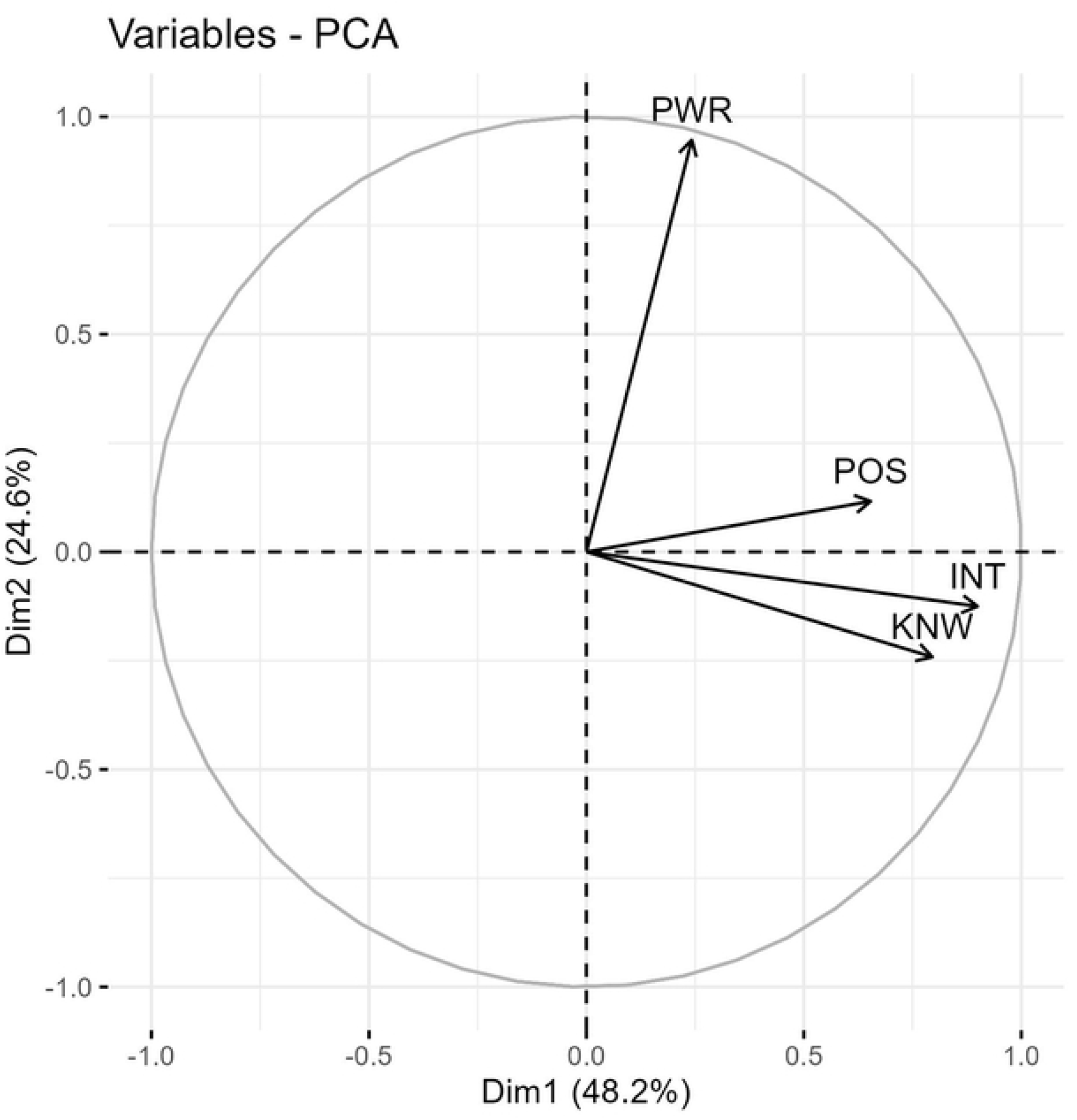
Principal Component Analysis (PCA) of the Four Characteristics: Influence (Y-axis) vs. Engagement (X-axis) The presented figure illustrates the results of a Principal Component Analysis (PCA) performed on the four characteristics: knowledge, interest, power, and position. The analysis generated two new components: Influence (represented on the Y-axis) and Engagement (represented on the X-axis). The PCA allows for the reduction of the multidimensional data into two principal components that capture the majority of the variance in the original dataset. The positioning of data points on the scatter plot indicates the level of influence (vertical axis) and engagement (horizontal axis) exhibited by each stakeholder. This representation enables a visual understanding of the relationships and patterns between the stakeholder characteristics and their impact on influence and engagement within the studied context.

The stakeholders who possess high interest are those who have either first-hand experience and knowledge about the RD ecosystem. INTO, PATO and RDS have extensive knowledge about the ecosystem. This acts as a base for their interest in policy development, implementation, and contributions to the ecosystem. The case is similar for other stakeholders such as TTNK, RCOM, and MNCO.

#### 3.5.3 Relationship between the stakeholder categories

A principal component analysis was conducted to investigate the inter-relationships among the four characteristics observed in the stakeholders. In Figure 4, the X and Y axes represent composite scores derived from the four characteristics. The angles formed by the three components (knowledge, interest, and position) are more acute, indicating a strong association among these variables. Additionally, the three components are primarily aligned with the X-axis. A new composite dimension termed ‘engagement’ is created by combining knowledge, interest, and position. Similarly, the dimension of ‘influence’ is derived from the power variable, which is predominantly aligned with the Y-axis.

Each stakeholder’s original scores on the four characteristics are transformed into two principal component scores and plotted on the two axes shown in Figure 5. Consequently, respondents 30 and 31 demonstrate high influence but only average engagement, while respondent 32 exhibits both high influence and greater engagement. Notably, all three stakeholders belong to the PMKR category. Conversely, other stakeholders in the PMKR category (27, 28, 29, 33, and 34) exhibit lower levels of influence and lesser engagement. Similarly, SHs 10 (LOCO), 7 (INTO), 21 (TTNK), 24 (RCOM), 20 (TTNK), 8 (INTO), 12 (LOCO), 37 (AHPR), 38 (AHPR) show high engagement, all of these stakeholders showed extensive knowledge and high interest.

**Figure 5.**
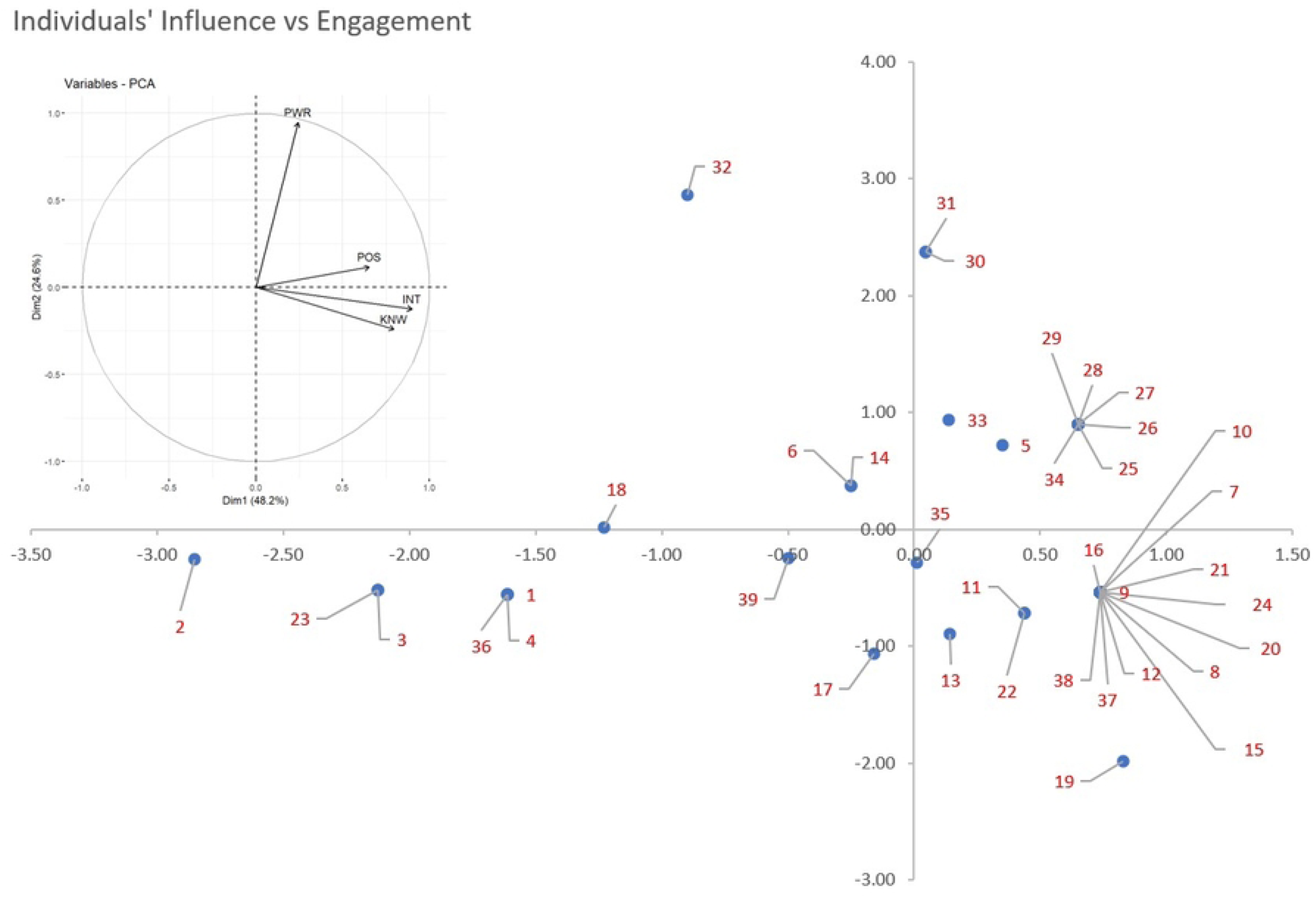
Principal component analysis and scatter plot of all the 39 stakeholders in the study. The scatter plot visually represents the positioning of each stakeholder in the reduced dimensional space derived from the PCA. The X and Y axes correspond to the influence and engagement obtained from the analysis. The scatter plot provides insights into the clustering, dispersion, and relationships among the stakeholders.

##### Challenges

- The interview also included a question on challenges and a diverse range of challenges were identified by different stakeholders which we summarize in Table 5.
- Most stakeholders identified lack of treatment in RDs in India, which they attribute to the healthcare system’s inadequate accessibility and availability of care. The situation is made worse by quack medicine, unqualified practitioners, and unreliable treatments. The government’s limited funding, delays in accessing approved treatments, and low awareness among the public and physicians further hinder effective management. Remote areas lack expertise, while general doctors show low interest and exposure to RDs. Inadequate research, public unwillingness to accept carrier status, lack of communication about policies, and gaps in healthcare coverage and system contribute to the challenges. Addressing these issues requires collaborative efforts, policy improvements, increased awareness, and better healthcare infrastructure.

##### Recommendations

During the interview process, the interviewers demonstrated their extensive knowledge and expertise by offering valuable recommendations for addressing the challenges that had been identified. Drawing from their many years of experience in their respective fields and their deep understanding of the RD ecosystem, they provided insightful solutions to tackle the identified issues. Their recommendations were rooted in a combination of theoretical knowledge and practical insights gained from navigating the complex landscape of RDs. These suggestions carried significant weight and provided a valuable roadmap for potential solutions, leveraging the interviewers’ rich expertise to guide future actions and decision-making processes.

1. Political will and funding crucial in fast-tracking RD policy activities - PATO, LOCO, TTNK, RDS
2. Improving accessibility of patients - PATO, TTNK
3. Improving access to clinical trials - MNCO, PATO, INTO
4. Giving importance to diagnosis irrespective of availability of treatment - MNCO, PATO, RDS
5. Creating specialty courses or training to personnel involved in RD ecosystem - PATO, TTNK, APHR
6. Measures to improve awareness regarding RDs - PATO, INTO, NRDS
7. Creating infrastructure in the form of treatment centers as well as the disabled friendly environment - PATO, PMKR
8. Creating collaborations with private and government entities - MNCO, LOCO
9. Inclusive environment for PAGs into the policymaking system - PATO
10. Improving research activities pertaining to RDs - RCOM, PATO
11. Creating a new screening process from the grass root level for RDs - PMKR, NRDS
12. Emotional and non-scientific support to be provided for patients and caretakers – PATO
13. Requirement for sensitization campaigns to promote empathy, acceptance, and support.

**Table 5:** Challenges in the RD ecosystem identified by different stakeholders.

| Challenges | Stakeholders who identified the challenges | Details |
| --- | --- | --- |
| Barriers to access to treatment | LOCO, PATO, MNCO, TTNK. | • Insufficient availability and accessibility of treatments for RDs within India's healthcare system.<br>• Limited options for patients resulting in delayed or inadequate care.<br>• Due to lack of approved treatment people find resort in unqualified practitioners and quacks with false claims |
| Regulatory challenges | PATO, TTNK, APHR, LOCO | • Challenges in obtaining timely access to treatments that have been approved by the U.S. Food and Drug Administration (FDA).<br>• Lengthy regulatory processes and delays in approval and availability of treatments in the Indian market. |
| Lack of funding | PATO, LOCO, TTNK, PMKR, INTO | • Limited financial support provided by the government for RD treatments.<br>• Insufficient allocation of resources for research, development, and access to therapies. |
| Lack of awareness and knowledge | NRDS, PATO, INTO, TTNK. | • Limited awareness, knowledge and understanding of RDs among the general public and healthcare professionals.<br>• Lack of knowledge leads to delays in diagnosis, misdiagnosis, referral, and appropriate management due to low awareness. |
| Stigma | PATO, RDS | • Reluctance among the public to acknowledge carrier status for RDs.<br>• Challenges in implementing carrier testing and counseling programs. |

## Discussion

RD specialists (RDS), International patient organizations (INTO), Local Companies (LOCO), Indian patient organizations (PATO), Think Tanks (TTNK), Research community (RCOM) and Multinational companies (MNCO)

Overall SH categories RDS, INTO, LOCO, PATO, TTNK, RCOM, and MNCO showed high knowledge, RDS, INTO, LOCO, PATO, TTNK, RCOM, and MNCO showed high interest, PMKR showed high power and AHPR, INTO, MNCO, PMKR and TTNK were strong supporters of the policy.

This study shows that there is a lack of awareness and knowledge about RDs among healthcare professionals who do not exclusively work in RDs viz: NRDS and AHPRs. These groups showed low scores in engagement in every aspect of knowledge, interest, and position which is concerning. Policies should therefore aim at gearing up awareness and training of all healthcare professionals and a strong reference network system needs to be developed that efficiently enables these professionals to identify patients and refer them to the right expert network.

PATOs are valuable groups as they score high on knowledge, interest and are capable of taking constructive critical stances in the policy making process as seen globally. However, they have scored low in power. It is imperative to encourage the active participation of RD patient organizations in the policy-making process related to RDs. To achieve this, it is crucial to formalize their involvement in the policy making process. The engagement of such organizations has proven to be highly beneficial on a global scale. For instance, in Europe, EURORDIS, a RD umbrella patient organization, plays a significant role in shaping policies and advocating for the needs of RD patients (10). Similarly, in the United States, NORD (National Organization for Rare Disorders) actively participates in implementing government agendas and influencing decision-making processes related to RDs (11).

Another important SH group is LOCO, they can be important players in driving innovation in RD treatment and management and make accessible treatments available to patients/ However, they exhibit low power, and more concerning is that MNCOs show higher scores on power than LOCO. This can have a significant negative impact on the local ecosystem where the needs of the LOCO may not be addressed and policies may end up going in the interest of more powerful MNCOs. In the end, this may result in making treatment inaccessible to the patients. Thus LOCOs need to be empowered to ensure a sustainable RD management and treatment ecosystem. We can take examples and inspiration towards steering the orphan drug approval process from countries like Brazil which has approved 21 orphan drugs and 31 clinical trials in 2019. This accomplishment demonstrates the regulator’s commitment to facilitating access to innovative treatments for RDs. Moreover, a recent regulatory development, the Resolution of the Collegiate Board (RDC) which became effective on June 1, 2020, signifies a significant step forward for Brazil. This resolution specifically addresses the registration process of advanced therapy products and outlines the regulations governing clinical trials involving these investigational medicines. Consequently, Brazil now possesses a robust regulatory framework to support the development and registration of cutting-edge high-technology products based on human cells and genes (12). RCOM and TTNK are another critical SHs who show high engagement and low influence. They play pivotal roles as important stakeholders in the RD ecosystem. Their contributions and expertise are instrumental in advancing our understanding of RDs, developing innovative treatments, and shaping policies and initiatives. The research community, comprising scientists, clinicians, and researchers, conducts groundbreaking studies to unravel the complexities of RDs. Their efforts contribute to the discovery of potential therapies, diagnostic tools, and interventions. By collaborating with patient organizations and healthcare professionals, they help bridge the gap between scientific discoveries and clinical practice, ultimately improving patient outcomes. TTKs, on the other hand, bring together experts from various disciplines to analyze and propose solutions to complex problems related to RDs. These organizations provide a platform for multidisciplinary discussions, policy analysis, and strategic planning. Their expertise aids in the formulation of evidence-based policies, guidelines, and initiatives that address the unique challenges faced by individuals living with RDs. The involvement of the research community and think tanks fosters a collaborative and multidimensional approach towards tackling RDs. Their contributions not only drive scientific advancements but also shape public health strategies, resource allocation, and advocacy efforts. By working in synergy with patient organizations, healthcare providers, and policymakers, they contribute to the development of a comprehensive and effective RD ecosystem.

Thus it is evident from our study that Indian RD policy decisions are largely shaped by PMKR, RDS, and MNCO. However, many important stakeholders such as PATO, RCOM, and TTNK have the potential to positively shape the RD agenda in the country based on their wide domain expertise and knowledge about the RD policy system. This highlights the need for a formal platform for inclusive participation of a wider network of individuals.

### Limitations of the study

- It was not possible to get representation from all stakeholder categories due to lack of response which is compensated by including media analysis for those stakeholder groups.
- Depth of stakeholders knowledge and engagement in one particular domain is not measured in the present study rather focus was on overall understanding of the RD ecosystem.

## Data Availability

Excerpts of the transcripts relevant to the transcript can be made available upon request but participants have not conseted to quote them in publication.

## Author contribution

MCC designed the project under the guidance of PNS. MCC conducted the interviews. MCC and JG independently analyzed the interview data, compared the extracted codes, and collated them. PN served as an advisor and provided guidance in project design and overall structuring. MCC and JG wrote the research paper and PNS reviewed it.

## Reflexivity Statement

MCC, as a RD policy researcher with six years of experience studying the RD ecosystem in India, brings extensive knowledge and familiarity with some of the stakeholders interviewed. This prior interaction may have shaped the understanding of their perspectives compared to stakeholders interviewed for the first time. PNS, with a background in public health unrelated to RDs, provided a neutral and unbiased perspective to the research. PNS intervened when any biases from MCC were noted, ensuring a balanced approach. JG, although lacking previous experience working with RDs, offered an unbiased opinion as a student of public health. The necessary skill sets acquired through his public health education enabled him to analyze the study objectively. By acknowledging these factors, we aim to enhance transparency and provide readers with insights into the potential subjectivity and influence of the researchers on the research process and findings.

## Ethical Approval

The study received approval from the Institutional Ethics Committee of the Institute of Public Health for the study number IEC-FR/01/2021 on 04/02/2021.

## Acknowledgement

We would like to extend our heartfelt appreciation to Manohar Rao for his invaluable contribution to the statistical analysis of our study. His expertise and insights have played a crucial role in shaping the outcomes and conclusions of our research. We thank the Department of Science and Technology, Government of India (DST) for providing the fellowship of MCC and PC. We also thank DST Center for Policy Research, and the Indian Institute of Science (IISc) and Institute of Public Health Bangalore for research support. We extend our sincere gratitude to Prof. T.A. Abinandanan, Coordinator for DST Centre for Policy Research IISc, for his unending support to the overall research work of MCC.

## Financial Disclosure

This study was funded by the DST/PRC/STI-PFP/2016 (IV Batch), dt:13.03.2020 DST STI Policy Fellowship awarded to MCC, Grant ID: DST/PRC/CPR/IISc-2023(G) (PCPM) Dt.31.03.2023.

The time contributions of PNS were supported by a CRC grant from DBT/ Wellcome Trust India Alliance, Grant ID: IA/CRC/20/1/600007 dt 24-09-2021 Host institution Ref. No. 06/2021

The funders had no role in study design, data collection and analysis, decision to publish, or preparation of the manuscript.

## Conflicts of interest

The authors declare no conflict of interest

## Consent

Written informed consent was collected from all the participants in the study.

## Competing interests

The authors declare that they have no competing interests.

## Notes

### Competing Interest Statement

The authors have declared no competing interest.

### Author Declarations

The study received approval from the Institutional Ethics Committee of the Institute of Public Health for study number IEC-FR/01/2021 on 04/02/2021.

